# Off-Label Dosing of Direct Oral Anticoagulants Among Inpatients with Atrial Fibrillation

**DOI:** 10.1101/2023.03.17.23287428

**Authors:** Amneet Sandhu, Lisa A. Kaltenbach, Karen Chiswell, Vijay Shimoga, Carmel Ashur, Abby Pribish, Gregg C. Fonarow, Jonathan P. Piccini, P. Michael Ho, Paul D. Varosy, Paul L. Hess

## Abstract

**Introduction:** Among patients hospitalized for atrial fibrillation (AF), the frequency of off-label direct oral anticoagulant (DOAC) dosing, associated factors, hospital-level variation, and temporal trends in contemporary clinical practice are unknown.

**Methods:** Using the Get With The Guidelines^®^ Atrial Fibrillation (GWTG-AF) registry, patients admitted from January 1^st^, 2014 to March 31^st^, 2020, and discharged on DOAC therapy were stratified according to receipt of underdosing, overdosing, or recommended dosing. Factors associated with off-label dosing were identified using logistic regression. Hospital-level variation and temporal trends were assessed.

**Results:** Of 22,470 patients prescribed a DOAC at discharge from hospitalization for AF (66% apixaban, 29% rivaroxaban, 5% dabigatran), underdosing occurred among 2006 (8.9%), overdosing among 511 (2.3%), and recommended dosing among 19953 (88.8%). Patient-related factors associated with off-label DOAC use included age (underdosing: OR 1.06 per 1-year increase [95% CI 1.06-1.07] and overdosing: OR 1.07 per 1-year increase [1.06-1.09]), dialysis dependence (underdosing: OR 5.50 [3.76-8.05] and overdosing: OR 5.47 [2.74-10.88]), female sex (overdosing: OR 0.79 [0.63-0.99]) and weight (overdosing: OR 0.96 per 1-Kg increase [0.95-1.00]). Across hospitals, the adjusted median odds ratio for off-label DOAC use was 1.45 [95% CI 1.34-1.65] (underdosing: 1.52 [1.39-1.76] and overdosing: 1.32 [1.20-1.84]), indicating significant hospital-level variation. Hospital characteristics associated with underdosing included West vs. Northeast location (OR: 1.55 [1.04-2.31]), rural vs. urban setting (OR: 0.48 [0.28-0.83]), and number of beds (<200 vs. 500+, OR: 1.95 [1.29-2.95]). Recommended dosing significantly increased over time (81.9% in 2014 to 90.9% in 2020, p<0.0001 for trend) with a corresponding decline in underdosing (14.4% in 2014 to 6.6% in 2020, p<0.0001 for trend) and overdosing (3.8% in 2014 to 2.5% in 2020, p=0.001 for trend).

**Conclusion:** One of 10 patients hospitalized for atrial fibrillation is discharged on off-label dosing of DOAC with significant variation across hospitals. While the proportion of patients receiving recommended dosing has significantly improved over time, opportunities to improve DOAC dosing persist.

## INTRODUCTION

Appropriately dosed direct oral anticoagulants (DOACs) reduce the risk of stroke and systemic embolism among select patients with atrial fibrillation (AF).^1-3^ The Food and Drug Administration (FDA) specified dosing derived from pivotal phase III trials^1-3^ based on factors inclusive of age, weight, kidney function, and concomitant medication use. Unfortunately, use of DOACs at doses not studied in the pivotal trials or recommended in FDA-labeling has been significant, affecting up to 12-20% of patients.^4-7^ Off-label DOAC dosing for AF has been observed in the outpatient setting and is associated with increased risk of cardiovascular hospitalization and all-cause mortality.^5,6^ Meta-analyses have also shown an increased bleeding risk with DOAC overdosing and higher stroke risk with DOAC underdosing.^8^

The degree to which off-label DOAC dosing occurs in patients hospitalized for AF is unknown. Hospitalizations often are associated with significant changes in health critical to prescription of an optimal DOAC dose, including weight, kidney function, and concurrent medication use. Temporal trends in off-label DOAC use and how use of these agents varies between hospitals is not known. Accordingly, using data from the Get With The Guidelines^®^-Atrial Fibrillation (GWTG-AFIB) registry, we sought to characterize (1) off-label DOAC dosing rates at discharge among patients requiring hospitalization for AF, (2) patient- and facility-level factors associated with off-label DOAC dosing and (3) temporal changes in the proportion of patients treated with off-label DOAC dosing.

## METHODS

### Data Source

The data used were collected by the American Heart Association’s Get With The Guidelines^®^-AFib registry. GWTG-AFIB registry was launched in 2013 as a prospective, national, observational initiative tracking hospital encounters for atrial fibrillation. The program and data elements of the GWTG-AFIB registry have been previously described.^9^ IQVIA (Parsippany, New Jersey) serves as the data collection and coordination center. A key objective of the GWTG program is to highlight national and institutional-level opportunities for quality improvement.

Each participating hospital received either human research approval to enroll cases without individual patient consent under the common rule, or a waiver of authorization and exemption from subsequent review by their institutional review board. The Duke Clinical Research Institute (Durham, NC) serves as the data analysis center and has an agreement to analyze the aggregate deidentified data for research purposes. The Institutional Review Board at Duke University Health approved this study. Participating sites were required to adhere to local regulatory and privacy procedures and obtain Institutional Review Board approval if needed. Institutional review board approval was granted to analyze limited data for research purposes.

### Study Population

The study population included patients who required hospital care for management of atrial fibrillation or atrial flutter and were discharged on a DOAC (apixaban, rivaroxaban or dabigatran between January 1^st^, 2014 and March 31^st^, 2020). Patient records that were (1) missing key demographic variables or medical history including age, sex, weight or history of atrial fibrillation, (2) missing discharge anticoagulant, dose, or frequency, (3) missing serum creatinine data at the time of discharge, (4) contraindications to DOAC or anticoagulant use or (5) document special circumstances (transition to comfort care or discharged against medical advice) or had missing destination after discharge were excluded (**Supplemental Figure 1**).

### Study Definition

The designation of “off-label” was defined as deviation from dosing specified by FDA package inserts and used in the seminal DOAC trials.^1-3^ They are based on age, weight, kidney function at discharge, and comorbid conditions such as need for dialysis (**Supplemental Table 1**).^10-12^ Recommended dosing of dabigatran varies by creatinine clearance (CrCl >30 ml/min = 150mg orally twice daily and CrCl 15-30 ml/min = 75 mg orally twice daily). For apixaban, recommended dosing is 5 mg twice daily. In the presence of any 2 of 3 factors comprised of age >80 years, weight <60 kg and a serum creatinine of >1.5 mg/dL, recommended dosing is 2.5mg orally twice daily. Recommended rivaroxaban dosing varies by creatinine clearance (>50 ml/min = 20mg orally once daily, 15-50 ml/min = 15mg orally once daily, and <15 ml/min = not recommended).

### Statistical Analysis

Patient characteristics were stratified by discharge DOAC dose characterized as underdosing, recommended dosing, or overdosing. Categorical variables were recorded as counts (percentages) and continuous variables reported as a median (Q1, Q3). Assessments of between-group differences were performed using Pearson χ^2^ or Fisher’s exact test as appropriate for the former and Kruskal-Wallis tests for the latter. In sensitivity analyses, rates of off-label dosing were assessed among patients with newly diagnosed AF, those were admitted on DOAC and discharged with a different prescription, and according to weight (< 60 kg, 60-120 kg, >120 kg or body mass index >40 Kg/m2).

To assess patient- and hospital-level factors associated with off-label dosing, overdosing was compared with recommended dosing and underdosing was compared with recommended dosing. In each case, a logistic regression model with stepwise selection was fitted using a significance of 0.10 to enter and remain in the model. Candidate variables were selected based on prior literature^5,6^ and clinical judgment which included demographics (age, sex, race and ethnicity), conditions affecting prior health (such as coronary artery disease, prior stroke or TIA, diabetes, hypertension, COPD, OSA, prior myocardial infarction, prior PCI, thyroid disease, prior hemorrhage, PVD, dialysis, liver disease, heart failure), other patient characteristics (such as left ventricular ejection fraction and history of other arrhythmias, smoking, insurance status), and hospital characteristics (region, academic, bedsize, rural). After variable selection, a random intercept for hospital to account for within hospital clustering was added. We assessed whether patient-or hospital-level factors play a larger role in inappropriate dosing using reference effect measures^13^, which compare patients in clusters at specified percentiles of the random effect of the distribution to patients with the same values for all measured covariates in a reference cluster. To make comparisons of site variability, variables were scaled such that odds ratios are comparable across variables. Whereas binary variables were dichotomized as in their original form, continuous variables were divided by 2*standard deviation (SD).

To characterize variation across hospitals, the percentage of patients with off-label dosing out of the total number of patients eligible for DOAC dosing was calculated for each hospital. Hospitals with <30 admissions in the study population were excluded. Hospital-level variation use was then graphically displayed using a caterpillar plot. To account for variation in the number of patients per site, a hierarchical logistic regression model with random intercepts for site was fitted. The model was then used to test whether variance components for site were greater than zero and to calculate the median odds ratio (MOR) between sites. The MOR can be interpreted as the median increase in odds of off-label dosing when an individual moves from a lower to a higher-risk hospital. It provides an estimate of the effect size of the hospital variation on the outcome of off-label DOAC dosing. A MOR >1.2 represents significant clinical variation.^14^

To describe temporal trends in percentage off-label DOAC dosing, the percentage of patients with off-label dosing was calculated by calendar quarter beginning in 2014. Trends in percentage of recommended dosing, underdosing, and overdosing over time was graphically displayed. To assess temporal trend significance, unadjusted and adjusted logistic regression models with random intercepts for site were fitted for overdosing versus recommended dosing and underdosing versus recommended dosing. The final clinical variables of the models described as well as random intercepts for site and time in quarters were included. The effect of quarterly trends of off-label DOAC dosing was estimated using odds ratios (95% confidence intervals).

## RESULTS

Among 22,470 patients discharged after a hospital encounter for AF, 2006 (8.9%) received a DOAC that was lower than the recommended dose, 19953 (88.8%) received a DOAC at recommended dosing, and 511 (2.3%) received a DOAC that was higher than the recommended dose. **Figure 1** displays underdosing, overdosing, and recommended dosing by DOAC type and overall.

**Figure 1:**
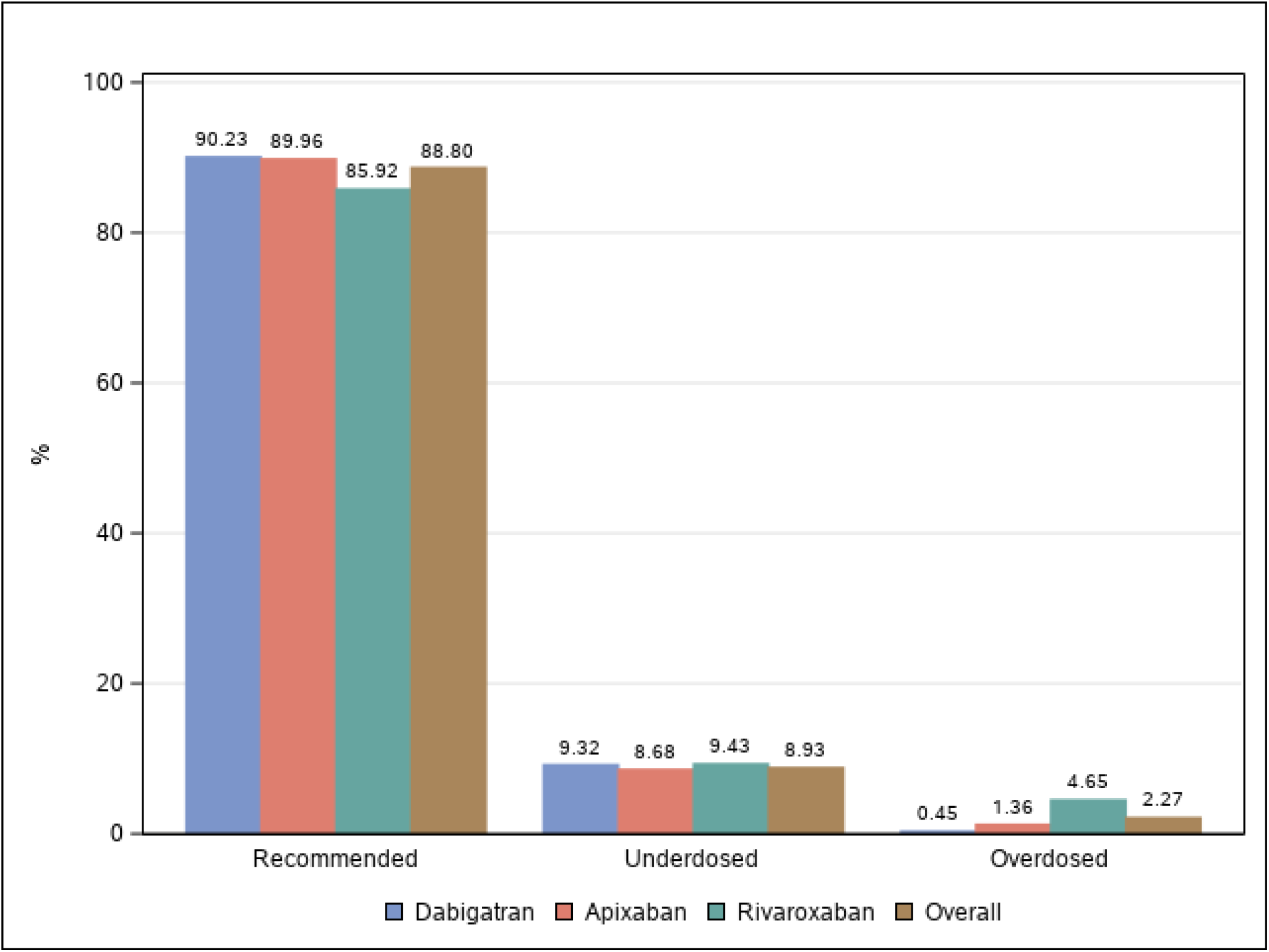
Rates of DOAC dosing at discharge in those hospitalized for atrial fibrillation, stratified by recommended dosing, underdosed or overdosed (percentages displayed).

**Table 1** displays patient-level data for the overall cohort and stratified by underdosed, recommended dosing and overdosed. In the overall population, the mean age was 70.1 +/-12.1 years old, 48.1% were female, and the mean body mass index was 31.2 +/-7.9 Kg/m^2^. The mean CHADS2Vasc score was 3.76 +/-1.75, 42.0% had paroxysmal AF and 37.6% had chronic kidney disease resulting in an eGFR <60 ml/min while 0.7% were on hemodialysis. Relative to those discharged on DOAC dosing consistent with FDA labeling, patients who received underdosed DOACs were older (77.0 +/-11.2 years vs. 69.1 +/-11.9), more commonly on dialysis (2.7% vs. 0.5%), more frequently had a prior hemorrhage (4.7% vs. 2.6%), and more frequently received care at hospitals located in non-rural settings (4.8% vs. 4.4%) or with less than 500 beds (43.4% vs. 33.8%). Relative to FDA-labeled use of DOACs on discharge, those who received overdosed DOACs were older (80.6 +/-7.8 years vs 69.1 +/-11.9), more frequently women (66.3% vs. 46.6%), usually had a lower body mass index (25.1 +/-5.8 vs. 31.5 +/-5.8), and more frequently were on dialysis (2.4% vs. 0.5%). In sensitivity analyses of rates of lower than recommended dose, recommended, and higher than recommended dose by new AF diagnosis, change in DOAC type (ie. from apixaban to rivaroxaban) during hospitalization or patient weight, findings were comparable to the primary analysis.

**Table 1:**
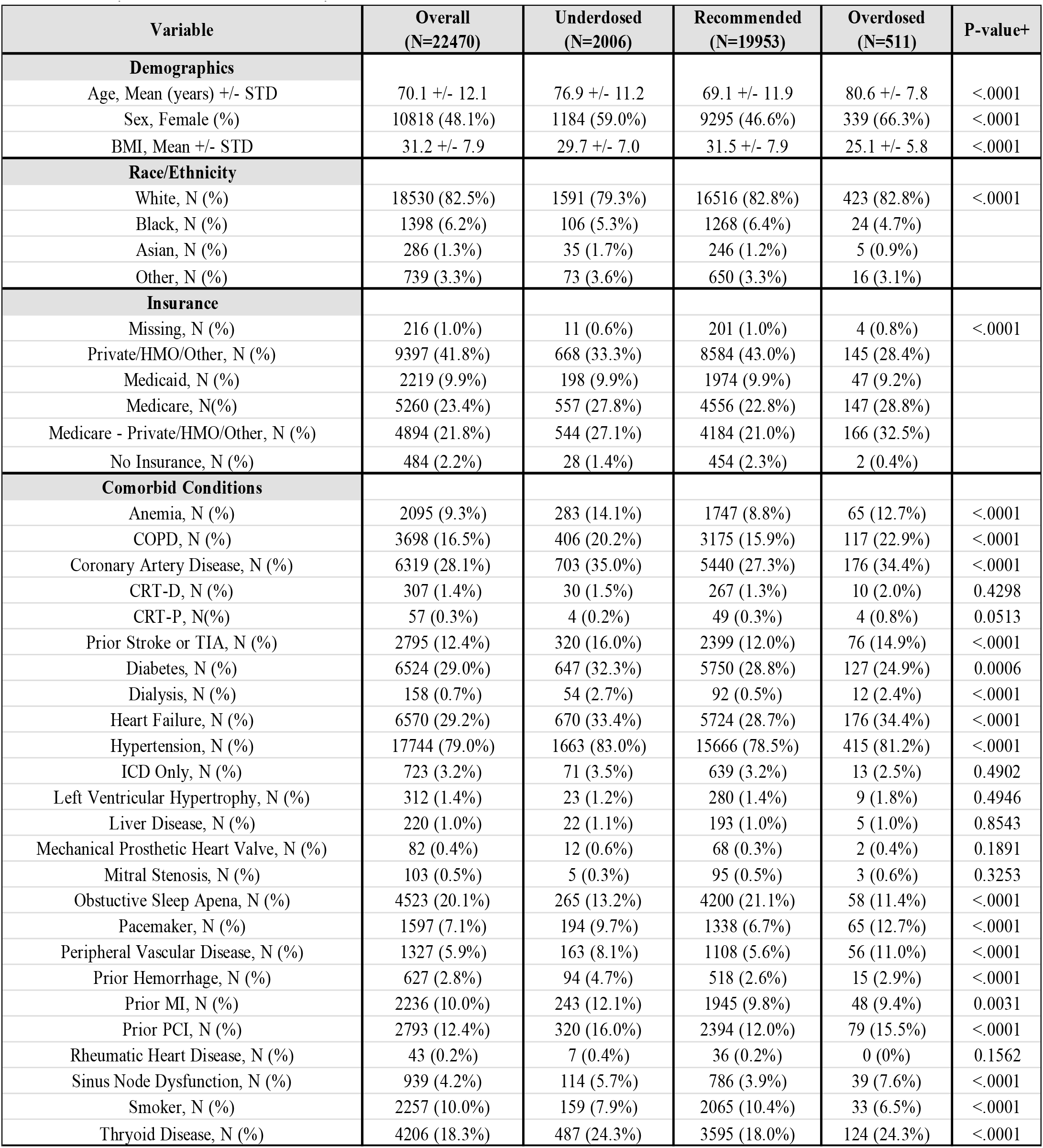
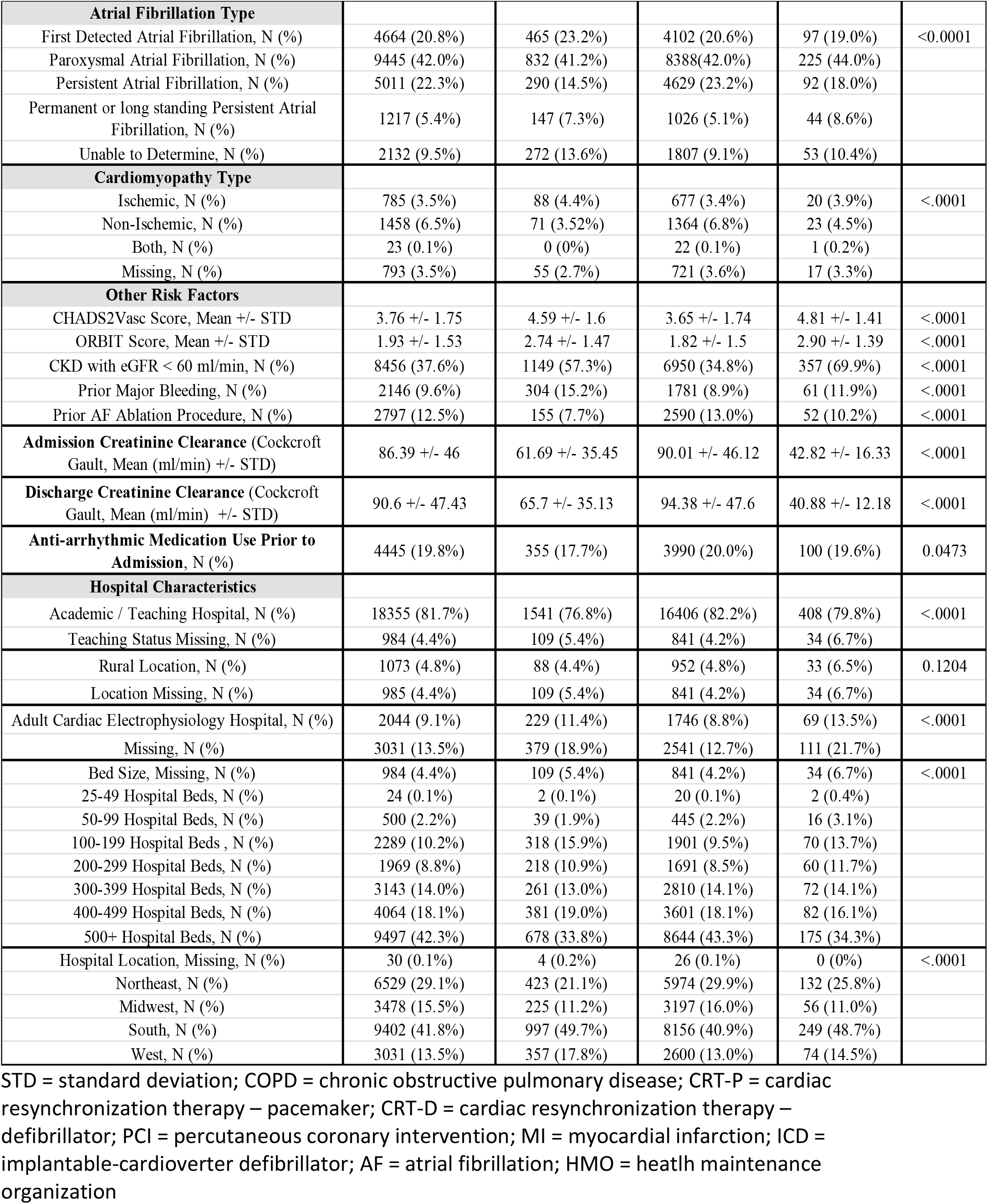
Summary of DOAC information, patient-level and hospital characteristics stratified by underdosed, recommended dose, overdosed and the overall cohort.

| Variable | Overall<br>(N=22470) | Underdosed<br>(N=2006) | Recommended<br>(N=19953) | Overdosed<br>(N=511) | P-value+ |
| --- | --- | --- | --- | --- | --- |
| **Demographics** |  |  |  |  |  |
| Age, Mean (years) +/- STD | 70.1 +/- 12.1 | 76.9 +/- 11.2 | 69.1 +/- 11.9 | 80.6 +/- 7.8 | <.0001 |
| Sex, Female (%) | 10818 (48.1%) | 1184 (59.0%) | 9295 (46.6%) | 339 (66.3%) | <.0001 |
| BMI, Mean +/- STD | 31.2 +/- 7.9 | 29.7 +/- 7.0 | 31.5 +/- 7.9 | 25.1 +/- 5.8 | <.0001 |
| **Race/Ethnicity** |  |  |  |  |  |
| White, N (%) | 18530 (82.5%) | 1591 (79.3%) | 16516 (82.8%) | 423 (82.8%) | <.0001 |
| Black, N (%) | 1398 (6.2%) | 106 (5.3%) | 1268 (6.4%) | 24 (4.7%) |  |
| Asian, N (%) | 286 (1.3%) | 35 (1.7%) | 246 (1.2%) | 5 (0.9%) |  |
| Other, N (%) | 739 (3.3%) | 73 (3.6%) | 650 (3.3%) | 16 (3.1%) |  |
| **Insurance** |  |  |  |  |  |
| Missing, N (%) | 216 (1.0%) | 11 (0.6%) | 201 (1.0%) | 4 (0.8%) | <.0001 |
| Private/HMO/Other, N (%) | 9397 (41.8%) | 668 (33.3%) | 8584 (43.0%) | 145 (28.4%) |  |
| Medicaid, N (%) | 2219 (9.9%) | 198 (9.9%) | 1974 (9.9%) | 47 (9.2%) |  |
| Medicare, N(%) | 5260 (23.4%) | 557 (27.8%) | 4556 (22.8%) | 147 (28.8%) |  |
| Medicare - Private/HMO/Other, N (%) | 4894 (21.8%) | 544 (27.1%) | 4184 (21.0%) | 166 (32.5%) |  |
| No Insurance, N (%) | 484 (2.2%) | 28 (1.4%) | 454 (2.3%) | 2 (0.4%) |  |
| **Comorbid Conditions** |  |  |  |  |  |
| Anemia, N (%) | 2095 (9.3%) | 283 (14.1%) | 1747 (8.8%) | 65 (12.7%) | <.0001 |
| COPD, N (%) | 3698 (16.5%) | 406 (20.2%) | 3175 (15.9%) | 117 (22.9%) | <.0001 |
| Coronary Artery Disease, N (%) | 6319 (28.1%) | 703 (35.0%) | 5440 (27.3%) | 176 (34.4%) | <.0001 |
| CRT-D, N (%) | 307 (1.4%) | 30 (1.5%) | 267 (1.3%) | 10 (2.0%) | 0.4298 |
| CRT-P, N(%) | 57 (0.3%) | 4 (0.2%) | 49 (0.3%) | 4 (0.8%) | 0.0513 |
| Prior Stroke or TIA, N (%) | 2795 (12.4%) | 320 (16.0%) | 2399 (12.0%) | 76 (14.9%) | <.0001 |
| Diabetes, N (%) | 6524 (29.0%) | 647 (32.3%) | 5750 (28.8%) | 127 (24.9%) | 0.0006 |
| Dialysis, N (%) | 158 (0.7%) | 54 (2.7%) | 92 (0.5%) | 12 (2.4%) | <.0001 |
| Heart Failure, N (%) | 6570 (29.2%) | 670 (33.4%) | 5724 (28.7%) | 176 (34.4%) | <.0001 |
| Hypertension, N (%) | 17744 (79.0%) | 1663 (83.0%) | 15666 (78.5%) | 415 (81.2%) | <.0001 |
| ICD Only, N (%) | 723 (3.2%) | 71 (3.5%) | 639 (3.2%) | 13 (2.5%) | 0.4902 |
| Left Ventricular Hypertrophy, N (%) | 312 (1.4%) | 23 (1.2%) | 280 (1.4%) | 9 (1.8%) | 0.4946 |
| Liver Disease, N (%) | 220 (1.0%) | 22 (1.1%) | 193 (1.0%) | 5 (1.0%) | 0.8543 |
| Mechanical Prosthetic Heart Valve, N (%) | 82 (0.4%) | 12 (0.6%) | 68 (0.3%) | 2 (0.4%) | 0.1891 |
| Mitral Stenosis, N (%) | 103 (0.5%) | 5 (0.3%) | 95 (0.5%) | 3 (0.6%) | 0.3253 |
| Obstructive Sleep Apnea, N (%) | 4523 (20.1%) | 265 (13.2%) | 4200 (21.1%) | 58 (11.4%) | <.0001 |
| Pacemaker, N (%) | 1597 (7.1%) | 194 (9.7%) | 1338 (6.7%) | 65 (12.7%) | <.0001 |
| Peripheral Vascular Disease, N (%) | 1327 (5.9%) | 163 (8.1%) | 1108 (5.6%) | 56 (11.0%) | <.0001 |
| Prior Hemorrhage, N (%) | 627 (2.8%) | 94 (4.7%) | 518 (2.6%) | 15 (2.9%) | <.0001 |
| Prior MI, N (%) | 2236 (10.0%) | 243 (12.1%) | 1945 (9.8%) | 48 (9.4%) | 0.0031 |
| Prior PCI, N (%) | 2793 (12.4%) | 320 (16.0%) | 2394 (12.0%) | 79 (15.5%) | <.0001 |
| Rheumatic Heart Disease, N (%) | 43 (0.2%) | 7 (0.4%) | 36 (0.2%) | 0 (0%) | 0.1562 |
| Sinus Node Dysfunction, N (%) | 939 (4.2%) | 114 (5.7%) | 786 (3.9%) | 39 (7.6%) | <.0001 |
| Smoker, N (%) | 2257 (10.0%) | 159 (7.9%) | 2065 (10.4%) | 33 (6.5%) | <.0001 |
| Thyroid Disease, N (%) | 4206 (18.3%) | 487 (24.3%) | 3595 (18.0%) | 124 (24.3%) | <.0001 |
| **Atrial Fibrillation Type** |  |  |  |  |  |
| First Detected Atrial Fibrillation, N (%) | 4664 (20.8%) | 465 (23.2%) | 4102 (20.6%) | 97 (19.0%) | <0.0001 |
| Paroxysmal Atrial Fibrillation, N (%) | 9445 (42.0%) | 832 (41.2%) | 8388 (42.0%) | 225 (44.0%) |  |
| Persistent Atrial Fibrillation, N (%) | 5011 (22.3%) | 290 (14.5%) | 4629 (23.2%) | 92 (18.0%) |  |
| Permanent or long standing Persistent Atrial Fibrillation, N (%) | 1217 (5.4%) | 147 (7.3%) | 1026 (5.1%) | 44 (8.6%) |  |
| Unable to Determine, N (%) | 2132 (9.5%) | 272 (13.6%) | 1807 (9.1%) | 53 (10.4%) |  |
| **Cardiomyopathy Type** |  |  |  |  |  |
| Ischemic, N (%) | 785 (3.5%) | 88 (4.4%) | 677 (3.4%) | 20 (3.9%) | <.0001 |
| Non-Ischemic, N (%) | 1458 (6.5%) | 71 (3.52%) | 1364 (6.8%) | 23 (4.5%) |  |
| Both, N (%) | 23 (0.1%) | 0 (0%) | 22 (0.1%) | 1 (0.2%) |  |
| Missing, N (%) | 793 (3.5%) | 55 (2.7%) | 721 (3.6%) | 17 (3.3%) |  |
| **Other Risk Factors** |  |  |  |  |  |
| CHADS2Vasc Score, Mean +/- STD | 3.76 +/- 1.75 | 4.59 +/- 1.6 | 3.65 +/- 1.74 | 4.81 +/- 1.41 | <.0001 |
| ORBIT Score, Mean +/- STD | 1.93 +/- 1.53 | 2.74 +/- 1.47 | 1.82 +/- 1.5 | 2.90 +/- 1.39 | <.0001 |
| CKD with eGFR < 60 ml/min, N (%) | 8456 (37.6%) | 1149 (57.3%) | 6950 (34.8%) | 357 (69.9%) | <.0001 |
| Prior Major Bleeding, N (%) | 2146 (9.6%) | 304 (15.2%) | 1781 (8.9%) | 61 (11.9%) | <.0001 |
| Prior AF Ablation Procedure, N (%) | 2797 (12.5%) | 155 (7.7%) | 2590 (13.0%) | 52 (10.2%) | <.0001 |
| **Admission Creatinine Clearance** (Cockcroft Gault, Mean (ml/min) +/- STD) | 86.39 +/- 46 | 61.69 +/- 35.45 | 90.01 +/- 46.12 | 42.82 +/- 16.33 | <.0001 |
| **Discharge Creatinine Clearance** (Cockcroft Gault, Mean (ml/min) +/- STD) | 90.6 +/- 47.43 | 65.7 +/- 35.13 | 94.38 +/- 47.6 | 40.88 +/- 12.18 | <.0001 |
| **Anti-arrhythmic Medication Use Prior to Admission, N (%)** | 4445 (19.8%) | 355 (17.7%) | 3990 (20.0%) | 100 (19.6%) | 0.0473 |
| **Hospital Characteristics** |  |  |  |  |  |
| Academic / Teaching Hospital, N (%) | 18355 (81.7%) | 1541 (76.8%) | 16406 (82.2%) | 408 (79.8%) | <.0001 |
| Teaching Status Missing, N (%) | 984 (4.4%) | 109 (5.4%) | 841 (4.2%) | 34 (6.7%) |  |
| Rural Location, N (%) | 1073 (4.8%) | 88 (4.4%) | 952 (4.8%) | 33 (6.5%) | 0.1204 |
| Location Missing, N (%) | 985 (4.4%) | 109 (5.4%) | 841 (4.2%) | 34 (6.7%) |  |
| Adult Cardiac Electrophysiology Hospital, N (%) | 2044 (9.1%) | 229 (11.4%) | 1746 (8.8%) | 69 (13.5%) | <.0001 |
| Missing, N (%) | 3031 (13.5%) | 379 (18.9%) | 2541 (12.7%) | 111 (21.7%) |  |
| Bed Size, Missing, N (%) | 984 (4.4%) | 109 (5.4%) | 841 (4.2%) | 34 (6.7%) | <.0001 |
| 25-49 Hospital Beds, N (%) | 24 (0.1%) | 2 (0.1%) | 20 (0.1%) | 2 (0.4%) |  |
| 50-99 Hospital Beds, N (%) | 500 (2.2%) | 39 (1.9%) | 445 (2.2%) | 16 (3.1%) |  |
| 100-199 Hospital Beds, N (%) | 2289 (10.2%) | 318 (15.9%) | 1901 (9.5%) | 70 (13.7%) |  |
| 200-299 Hospital Beds, N (%) | 1969 (8.8%) | 218 (10.9%) | 1691 (8.5%) | 60 (11.7%) |  |
| 300-399 Hospital Beds, N (%) | 3143 (14.0%) | 261 (13.0%) | 2810 (14.1%) | 72 (14.1%) |  |
| 400-499 Hospital Beds, N (%) | 4064 (18.1%) | 381 (19.0%) | 3601 (18.1%) | 82 (16.1%) |  |
| 500+ Hospital Beds, N (%) | 9497 (42.3%) | 678 (33.8%) | 8644 (43.3%) | 175 (34.3%) |  |
| Hospital Location, Missing, N (%) | 30 (0.1%) | 4 (0.2%) | 26 (0.1%) | 0 (0%) | <.0001 |
| Northeast, N (%) | 6529 (29.1%) | 423 (21.1%) | 5974 (29.9%) | 132 (25.8%) |  |
| Midwest, N (%) | 3478 (15.5%) | 225 (11.2%) | 3197 (16.0%) | 56 (11.0%) |  |
| South, N (%) | 9402 (41.8%) | 997 (49.7%) | 8156 (40.9%) | 249 (48.7%) |  |
| West, N (%) | 3031 (13.5%) | 357 (17.8%) | 2600 (13.0%) | 74 (14.5%) |  |

STD = standard deviation; COPD = chronic obstructive pulmonary disease; CRT-P = cardiac resynchronization therapy – pacemaker; CRT-D = cardiac resynchronization therapy – defibrillator; PCI = percutaneous coronary intervention; MI = myocardial infarction; ICD = implantable-cardioverter defibrillator; AF = atrial fibrillation; HMO = health maintenance organization

**Figures 2** displays factors associated with overdosing and underdosing relative to recommended dosing as well as measures of variation across hospitals. In multivariable modeling, higher rates of underdosing were associated with patient-level factors such as older age, dialysis dependence, and prior hemorrhage and hospital-level factors such as Western and urban location as well as servicing relatively few beds. Across hospitals, the reference effect measure for random site variation of receipt of underdosed DOAC at discharge (90^th^ percentile in comparison to median hospital) was OR 1.75 (95% CI 1.54-2.00). Higher rates of overdosing were associated with patient-level factors such as older age and dialysis dependence. Hospital-level factors were not significantly contributory. Across hospitals, the reference effect measure for random site variation of receipt of an overdosed DOAC at discharge (90^th^ percentile in comparison to median hospital) was OR 1.46 (95% CI 1.29-1.65).

**Figure 2:**
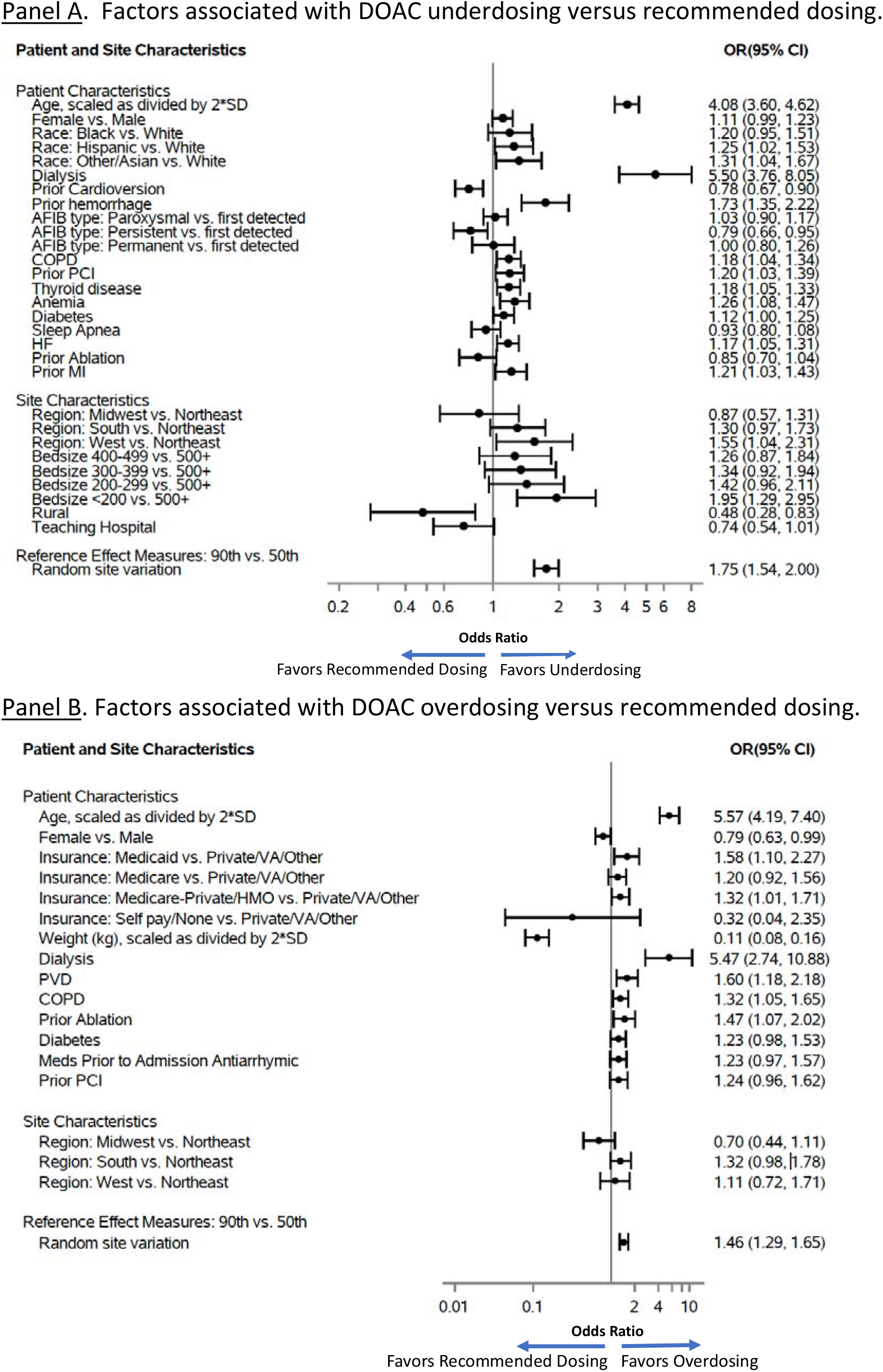
Factors associated with off-label dosing (Panel A: Underdosing and Panel B: Overdosing).

Across participating sites, the median observed percent of off-label DOAC use was 10.9% (IQR 6.8 – 15.9%). **Figure 3** displays hospital-level variation in the rate of off-label DOAC use in addition to hospital-level variation in rates of underdosing and overdosing. The overall adjusted MOR for off-label DOAC use across hospitals was 1.45 (95% CI 1.34-1.65) [REM or random effects model range 0.47-2.14], indicating significant variation across sites. The adjusted MOR for underdosing was 1.52 (95% CI 1.39-1.76) [REM range 0.42-2.36] and overdosing 1.32 (1.20-1.84) [REM range 0.56-1.78]. **Table 2** shows that in random effects models, while patient factors contributed more to variability than facility-level factors, facility-level factors were nonetheless significantly contributory to both underdosing and overdosing. **Figure 4** shows temporal trends in the rates of recommended dosing, underdosing, and overdosing of DOACs. There was a significant increase in recommended dosing from 81.9% in 2014 to 90.9% in 2020, p <0.0001 for trend. There was a significant decline in those receiving underdosing (14.4% in 2014 to 6.6% in 2020, p<0.0001 for trend) and overdosing (3.8% in 2014 to 2.5% in 2020, p=0.001 for trend) dosing over the study period.

**Table 2:** Contribution to Hospital-Level Variation by Groups of Factors: All Factors, Patient-Level Factors, and Hospital-Level Factors

|  |  | Reference Effect Measures* ranges and percentiles |  |  |
| --- | --- | --- | --- | --- |
| Outcome | Variables | Range | 10<sup>th</sup> | 90<sup>th</sup> |
| Underdosed | All factors | [0.147, 5.496] | 0.289 | 3.224 |
|  | Patient factors | [0.172, 4.325] | 0.328 | 2.826 |
|  | Hospital factors | [0.669, 2.159] | 0.77 | 1.698 |
| Overdosed | All factors | [0.017, 13.797] | 0.082 | 6.606 |
|  | Patient factors | [0.017, 12.794] | 0.085 | 6.34 |
|  | Hospital factors | [0.630, 1.187] | 0.63 | 1.187 |

\*Compares patients clustered within hospitals at specified percentiles of random effect distributions to similar patients in a reference, median hospital. Wider ranges indicate larger contributions to overall variation in outcome from variables.

**Figure 3:**
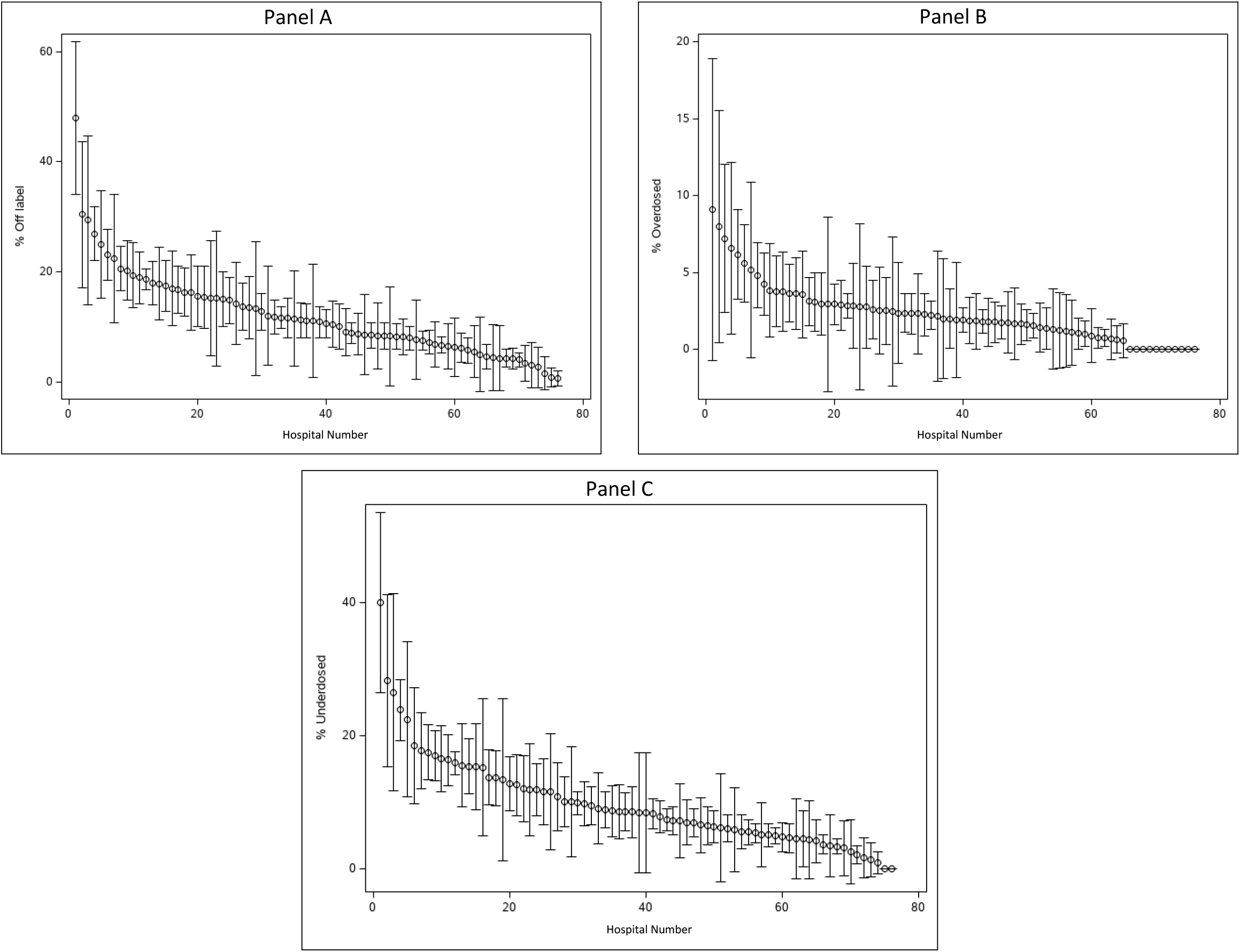
Facility-level variation in the rate of off-label DOAC use (Panel A), overdosing (Panel B) and underdosing (panel C) at discharge

**Figure 4:**
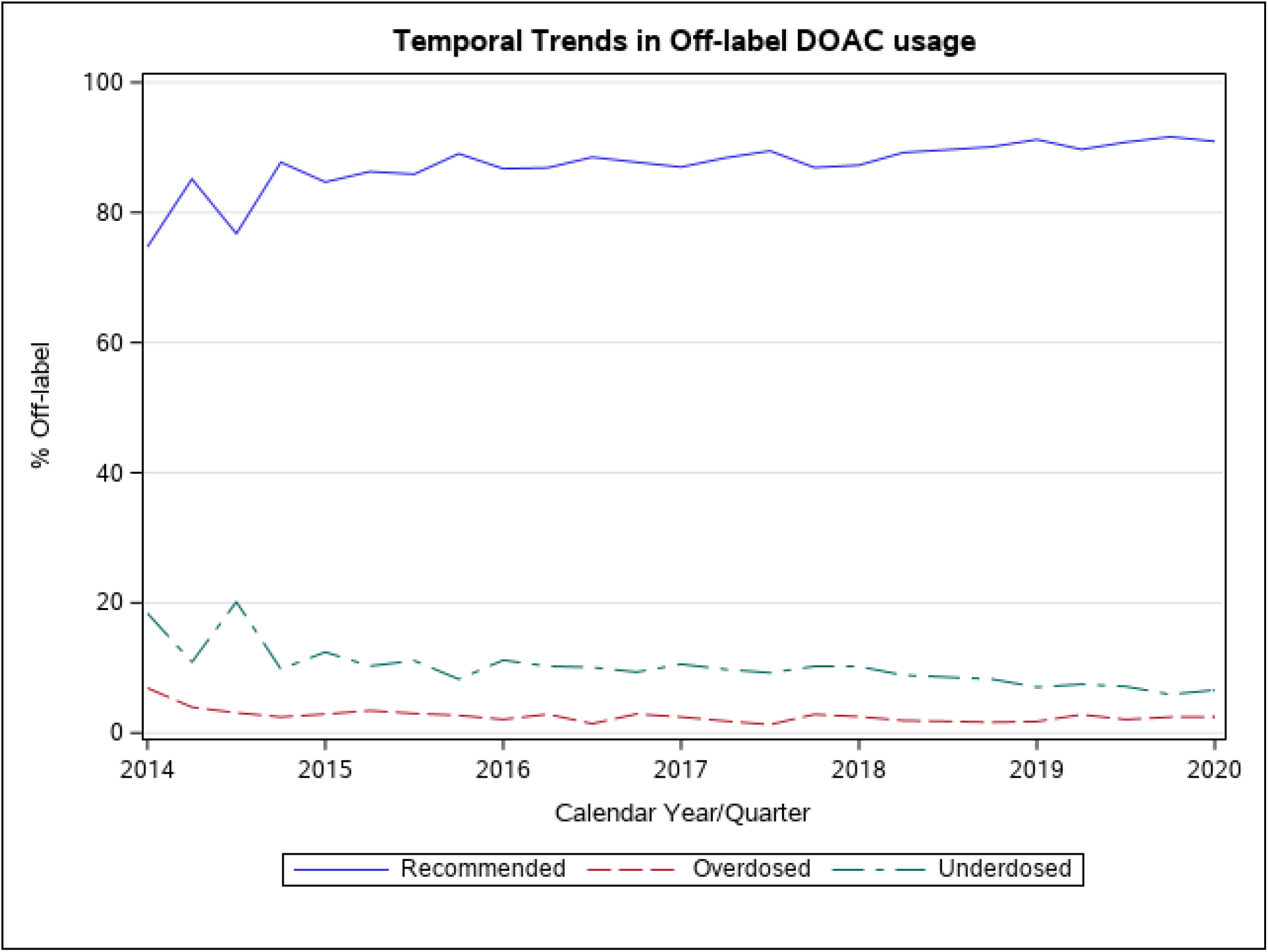
Trends, from 2014-2020, in the rates of recommended, under and overdosing of DOACs.

## DISCUSSION

In this nationwide analysis of more than 22,000 patients hospitalized for care of atrial fibrillation and discharged on DOACs, there are three key findings. First, 1 of 10 patients hospitalized for AF receive under or overdosed DOACs. Second, significant hospital-level variation exists with regards to use of off-label DOAC dose, with the greatest opportunity for future improvement in hospitals that are Western, urban, or of comparatively small size. Third, over the study period, rates of recommended DOAC dosing increased and off-label use decreased. These results characterize favorable national trends in DOAC use while identifying continued opportunities to improve safe and appropriate DOAC dosing at hospital discharge.

Prior work has analyzed rates of off-label DOAC dose in ambulatory outpatients (9.4% are underdosed, 3.4% overdosed and 87% per recommendation) correlating adverse cardiovascular or bleeding events in those who received off-label dosing.^6^ Our work extends that of prior analyses by focusing on patients hospitalized with AF and reveals rates of off-label use (8.9% underdosed, 2.3% overdosed) similar to that seen among outpatients.^6,15^ Encouragingly, rates of recommended dosing remain relatively high and are comparable to prior, smaller analyses in patients with atrial fibrillation warranting long-term anticoagulation.^16^ Consistent with prior, smaller studies, we found that several patient characteristics are more common in those treated with an off-label dose, including older age, weight, and dialysis dependence.^6,17^ The DOACs studied (apixaban, rivaroxaban and dabigatran) account for the majority of DOAC use in the United States.^18,19^ Broadly, these data provide opportunities to address this quality gap, focusing on patient profiles at-risk for off-label DOACs dose, risk of over or underdosing based on type of DOAC utilized. These profiles may prove useful both at the time of hospital discharge as well as during post-hospitalization follow-up clinical encounters.

Significant hospital-level variation of off-label DOACs use exists after accounting for measured variables. This finding suggests unmeasured aspects of site-level care may account for a significant proportion of hospital-variation. Such aspects may include variability in formalized structure surrounding quality improvement such as that recommended for dedicated AF Centers of Excellence.^20^ In this context, system-level quality improvement efforts may prove fruitful, primarily focusing on reducing rates of underdosing. This may be achieved with the the development of team-based, integrated clinical care pathways developed by relevant stakeholders, including pharmacy, nursing, hospital medicine, and cardiology. Key components may include the establishment of pre-discharge medication review processes, enhanced clinical decision support with automated dosing checks accounting for key comorbidities, medications, and up-to-date laboratory values embedded in the electronic medical record, and close outpatient follow-up attentive to the importance of appropriate DOAC dosing.

Nationwide improvement in rates of recommended DOAC use and decline in the use of off-label use suggest there may already be some level of recognition of the importance of appropriately-dosed anticoagulation. Nonetheless, the presence of a significant, persistent gap and heterogeneity in performance across hospitals underscores the need for continued, focused mitigation efforts. Endeavors may include not only system-level quality improvement programs described above but also benchmarking of DOAC dosing and the development of performance measures. Benchmarking such as that provided by quality improvement registries like GWTG-AF is likely an effective feedback mechanism to stimulate improvement. In addition, establishment of provider- and facility-level AF performance measures related to appropriate AF dosing may also prove to be an effective a feedback mechanism and policy incentive for continued quality improvement.

Limitations of our work include analysis of hospitals participating in the GWTG-AF Registry, which may select for hospitals choosing to be involved with quality improvement work. Though missing data was the major contributor to patient exclusion, the primary sample size of >20,000 patients allow for meaningful analyses and represents a much greater sample in comparison to other works evaluating off-label dosing. Factors considered by clinicians such as frailty may influence DOAC dosing and and yet are not captured in the GWTG-AF registry. The intent of our work was to evaluate rates of and factors associated with recommended and off-label DOAC dose in those hospitalized for atrial fibrillation, and as such whether off-label DOAC dosing persisted after discharge was not evaluated. However, this study represents (1) the largest work, to date, analyzing contemporary DOAC dosing and (2) the first to evaluate patients hospitalized for AF and discharged on DOAC therapy.

## CONCLUSIONS

One in 10 patients hospitalized for AF are discharged on off-label doses of DOAC, with significant variation across hospitals. Over time, rates of underdosing and overdosing declined while the rate of recommended DOAC dosing increased. Owing to persistently elevated rates of off-label DOAC dosing, quality-improvement efforts should be considered.

## Data Availability

De-identified data will be made available upon request from the corresponding author, pending permission from the AHA-GWTG program.

## Appendix

**Supplemental Table 1:** Baseline dosing recommendation and dosing adjustments for commonly prescribed DOACs in the United States.

|  | Baseline Dosing Recommendation | Dosing Adjustments |
| --- | --- | --- |
| Dabigatran | 150mg PO BID for CrCl >30 ml/min | 1. CrCl 30-50 ml/min = if receiving concomitant dronedarone, reduce to 75mg PO BID<br>2. 15-30 ml/min = 75mg PO BID unless on dronedarone, then avoid concurrent use<br>3. CrCl <15 ml/min = Not Recommended |
| Apixaban | 5mg PO BID | 1. 2.5mg PO BID recommended if any 2 of 3 factors present: age ≥80 years old, weight ≤60 kg or Creatinine ≥1.5 mg/dL<br>2. If on dialysis: 5mg PO BID, reduce to 2.5mg BID if age ≥80 years or weight ≤60 Kg |
| Rivaroxaban | 20mg PO qDay for CrCl >50 ml/min | 1. CrCl 15-50 ml/min = 15mg PO qDay<br>2. CrCl <15 ml/min = Not Recommended |

**Supplemental Figure 1:**
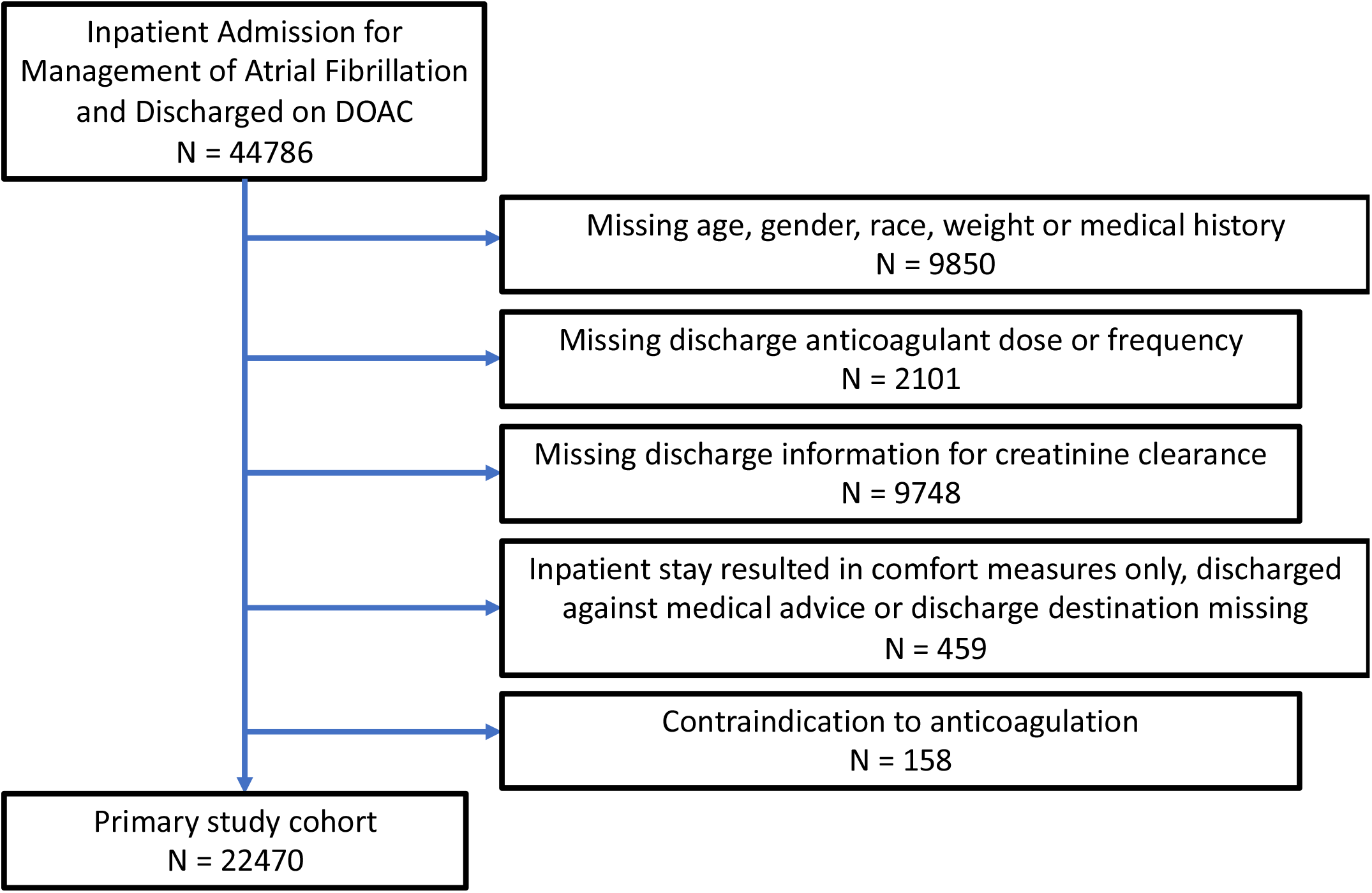
Attrition plot deriving the primary cohort of interest.

